# RNA-Based COVID-19 Vaccine BNT162b2 Selected for a Pivotal Efficacy Study

**DOI:** 10.1101/2020.08.17.20176651

**Authors:** Edward E. Walsh, Robert Frenck, Ann R. Falsey, Nicholas Kitchin, Judith Absalon, Alejandra Gurtman, Stephen Lockhart, Kathleen Neuzil, Mark J. Mulligan, Ruth Bailey, Kena A. Swanson, Ping Li, Kenneth Koury, Warren Kalina, David Cooper, Camila Fontes-Garfias, Pei-Yong Shi, Özlem Türeci, Kristin R. Tompkins, Kirsten E. Lyke, Vanessa Raabe, Philip R. Dormitzer, Kathrin U. Jansen, Uğur Şahin, William C. Gruber

**Affiliations:** University of Rochester and Rochester General Hospital, Rochester, NY; Cincinnati Children’s Hospital, Cincinnati, OH; Vaccine Research and Development, Pfizer Inc; Hurley, UK; Pearl River, NY; Collegeville, PA; University of Maryland School of Medicine, Center for Vaccine Development and Global Health, Baltimore, MD; New York University Langone Vaccine Center and New York University Grossman School of Medicine, New York, NY; University of Texas Medical Branch, Galveston, TX; BioNTech, Mainz, Germany

**Keywords:** COVID-19, SARS-CoV-2, vaccine, mRNA, adults

## Abstract

**Background:** Severe acute respiratory syndrome coronavirus 2 (SARS-CoV-2) infections and the resulting disease, coronavirus disease 2019 (COVID-19), have spread to millions of people globally. Multiple vaccine candidates are under development, but no vaccine is currently available.

**Methods:** Healthy adults 18–55 and 65–85 years of age were randomized in an ongoing, placebo-controlled, observer-blinded dose-escalation study to receive 2 doses at 21-day intervals of placebo or either of 2 lipid nanoparticle–formulated, nucleoside-modified RNA vaccine candidates: BNT162b1, which encodes a secreted trimerized SARS-CoV-2 receptor-binding domain, or BNT162b2, which encodes a prefusion stabilized membrane-anchored SARS-CoV-2 full-length spike. In each of 13 groups of 15 participants, 12 received vaccine and 3 received placebo. Groups were distinguished by vaccine candidate, age of participant, and vaccine dose level. Interim safety and immunogenicity data of BNT162b1 in younger adults have been reported previously from US and German trials. We now present additional safety and immunogenicity data from the US Phase 1 trial that supported selection of the vaccine candidate advanced to a pivotal Phase 2/3 safety and efficacy evaluation.

**Results:** In both younger and older adults, the 2 vaccine candidates elicited similar dose-dependent SARS-CoV-2-neutralizing geometric mean titers (GMTs), comparable to or higher than the GMT of a panel of SARS-CoV-2 convalescent sera. BNT162b2 was associated with less systemic reactogenicity, particularly in older adults.

**Conclusion:** These results support selection of the BNT162b2 vaccine candidate for Phase 2/3 large-scale safety and efficacy evaluation, currently underway.

## INTRODUCTION

Since the first coronavirus disease 2019 (COVID-19) cases in Wuhan, China, in December 2019, pandemic illness has spread to millions of people globally. No COVID-19 vaccines are currently available, and they are urgently needed to combat escalating cases and deaths worldwide.^1^

In response, BioNTech and Pfizer launched an unprecedented and coordinated program to compare 4 RNA-based COVID-19 pandemic vaccine candidates in umbrella-type clinical studies conducted in Germany (BNT162-01) and the US (C4591001). The program was designed to support the selection of a single vaccine candidate and dose level for a pivotal global safety and efficacy trial. Based on initial clinical trial results in Germany,^2^ 2 lipid nanoparticle–formulated,^3^ nucleoside-modified RNA (modRNA)^4^ vaccine candidates were evaluated in the US Phase 1 portion of the trial.^5^ One of these, BNT162b1, encodes the SARS-CoV-2 receptor-binding domain (RBD), trimerized by the addition of a T4 fibritin foldon domain to increase its immunogenicity through multivalent display.^6,7,8^ The other, BNT162b2, encodes the SARS-CoV-2 full-length spike, modified by 2 proline mutations (P2 S) to lock it in the prefusion conformation^9^ to increase its potential to elicit virus-neutralizing antibodies.^10^

Previous publications have described assessment of BNT162b1 in 18–55 year old healthy adults at multiple dose levels.^2,5^ Those studies indicated that well-tolerated dose levels of BNT162b1 efficiently elicited high titer, broad serum neutralizing responses, T_H_1 phenotype CD4^+^ T helper cell responses, and strong interferon γ (IFN-γ) and interleukin-2 (IL-2) producing CD8+ cytotoxic T-cell responses. This ability to elicit both humoral and cell-mediated antiviral mechanisms makes BNT162b1 a promising vaccine candidate.

Here we report the full set of safety and immunogenicity data from the Phase 1 portion of an ongoing randomized, placebo-controlled, observer-blinded dose-escalation US trial that was used to select the final vaccine candidate, BNT162b2, as well as comparison of the safety and immunogenicity of both vaccine candidates (ClinicalTrials.gov identifier: NCT04368728). These data include evaluation of 10-μg, 20-μg, and 30-μg dose levels of BNT162b1 in 65–85 year old adults and of an additional 20-μg dose level in 18–55 year old adults. In addition, the safety, tolerability, and immunogenicity of BNT162b2 in both younger and older adults are compared to those of BNT162b1 at 10-μg, 20-μg, and 30-μg dose levels.

## METHODS

### Trial Objectives

Safety and immunogenicity of varying dose levels of BNT162b1 and BNT162b2.

### Trial Participants

Healthy adults 18–55 or 65–85 years of age were eligible for inclusion. Key exclusion criteria included: known infection with HIV, HCV, or HBV; immunocompromised; history of autoimmune disease; previous clinical or microbiological diagnosis of COVID-19; receipt of medications intended to prevent COVID-19; prior coronavirus vaccination; a positive test for SARS-CoV-2 IgM and/or IgG at the screening visit; and a SARS-CoV-2 nucleic acid amplification test (NAAT)-positive nasal swab within 24 hours before study vaccination.

### Trial Procedures

Using an interactive web-based response technology system, study participants were randomly assigned to vaccine groups defined by vaccine candidate, dose level, and age range. Participants received two 0.5-mL injections into the deltoid of either BNT162b1, BNT162b2, or placebo, 21 days apart. The first 5 participants in each new dose level or age group were observed for 4 hours after vaccination to identify immediate adverse events (AEs). All other participants were observed for 30 minutes. Blood was collected for safety and/or immunogenicity assessments.

### Safety

Primary endpoints presented include: the proportions of participants with solicited local reactions, systemic events, and use of antipyretic and/or pain medication within 7 days after vaccination recorded in an electronic diary; unsolicited AEs and serious adverse events (SAEs) from Dose 1 through 1 month after Dose 2; clinical laboratory abnormalities 1 and 7 days after vaccination; and grading shifts in laboratory assessments between baseline and 1 and 7 days after Dose 1 and between 2 and 7 days after Dose 2. There were protocol-specified safety stopping rules for all sentinel-cohort participants. An internal review committee and an external data monitoring committee reviewed all safety data.

### Immunogenicity

Immunogenicity assessments (SARS-CoV-2 serum neutralization assay and RBD-binding or S1-binding IgG direct Luminex immunoassays) were assessed before vaccination, at 7 and 21 days after Dose 1, and 7 days (Day 21) and 14 days (Day 35) after Dose 2. The neutralization assay used a previously described strain of SARS-CoV-2 (USA_WA1/2020) that had been rescued by reverse genetics and engineered by the insertion of an mNeonGreen gene into open reading frame 7 of the viral genome.^11,12^ The 50% neutralization titer (NT_50_) was reported as the interpolated reciprocal of the dilution yielding a 50% reduction in fluorescent viral foci. One serum sample from the 30 μg BNT162b2 18–55 year old group, drawn 3 days after Dose 2 rather than 6 to 8 days after Dose 2 (as specified in the protocol), was excluded from the reported immunogenicity analysis. No other samples were reported to be drawn outside of the protocol-specified window.

Immunogenicity data from a human convalescent serum panel are included as a benchmark. Thirty-eight human SARS-CoV-2 infection/COVID-19 convalescent sera were drawn from donors 18 to 83 years of age, at least 14 days after a PCR-confirmed diagnosis and after symptom resolution. Neutralizing GMTs in subgroups of the donors were as follows: symptomatic infections – 90 (n=35); asymptomatic infections – 156 (n=3); hospitalized – 618 (n=1). The sera were obtained from Sanguine Biosciences (Sherman Oaks, CA), the MT Group (Van Nuys, CA), and Pfizer Occupational Health and Wellness (Pearl River, NY).

### Statistical Analysis

In this report, safety and immunogenicity analyses are descriptive, and the sample size was not based on statistical hypothesis testing. Safety analyses are presented as counts, percentages, and associated Clopper-Pearson 95% confidence intervals (CIs) for local reactions, systemic events, and any AEs after vaccination according to MedDRA terms for each vaccine group. Summary statistics are provided for abnormal laboratory values and grading shifts.

Immunogenicity analyses of SARS-CoV-2 serum neutralizing titers, S1- and RBD-IgG binding concentrations, GMTs, and geometric mean concentrations (GMCs) were computed along with associated 95% CIs. The GMTs/GMCs were calculated as the mean of the assay results after making the logarithm transformation and then exponentiating the mean to express results on the original scale. Two-sided 95% CIs were obtained by taking log transforms of titers or concentrations, calculating the 95% CI with reference to Student’s t-distribution, and then exponentiating the confidence limits.

## RESULTS

### Disposition and Demographics

Between 04 May 2020 and 22 June 2020, 332 healthy males and nonpregnant females at 4 US sites (2 sites per vaccine candidate) were screened: 195 were randomized to 13 groups of 15 participants (12 received vaccine and 3 placebo per group) (Figure 1). Groups of 18–55 year old and 65–85 year old participants received 10-μg, 20-μg, or 30-μg dose levels of BNT162b1 or BNT162b2 on a 2-dose schedule, 21 days apart. One group of 18–55 year old participants received 1 dose of 100 μg BNT162b1 or placebo. Overall, participants were predominantly white (67%-100%) and non-Hispanic/non-Latinx (92%-100%) (Table 1). More older women than older men participated. The median age of younger participants was 35–37 years and of older participants was 68–69 years, depending on vaccine candidate.

**Figure 1.**
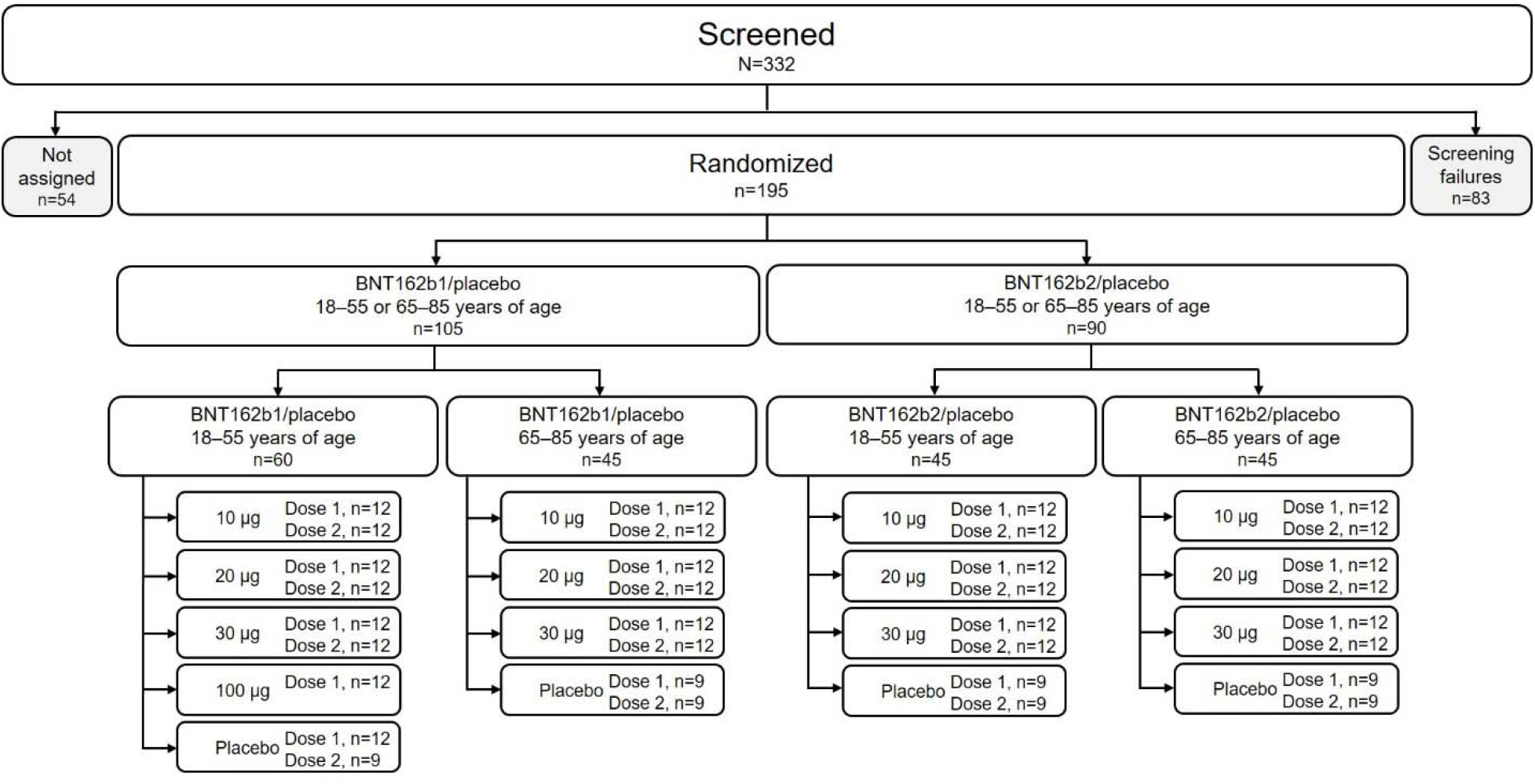
| Disposition of participants. Participants not assigned were screened but not randomized because enrollment had closed.

**Table 1.**
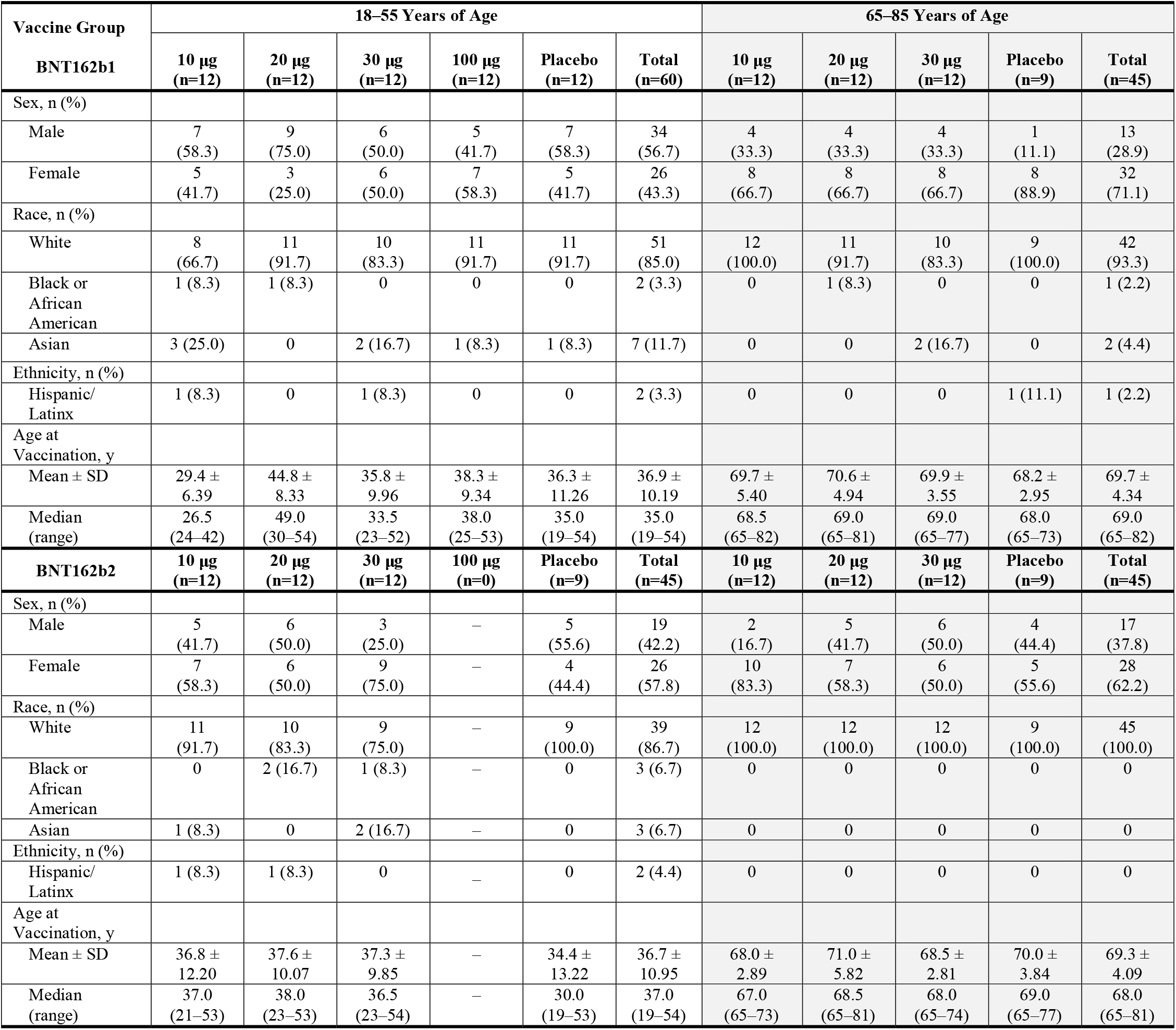
| Participant demographics for BNT162b1 and BNT162b2.

### Safety

#### Reactogenicity

##### Local reactions

Participants 18–55 years old who received 10 or 30 μg BNT162b1 reported mild to moderate local reactions, primarily pain at the injection site, within 7 days after an injection, which were more frequent after Dose 2.^2, 5^ In 65–85 year olds, BNT162b1 elicited similar but milder local reactions, with mild to moderate injection site pain reported by 92% after Dose 1 and 75% after Dose 2 (Figure 2). A similar pattern was observed after vaccination with BNT162b2. No older adult who received BNT162b2 reported redness or swelling. No participant who received either BNT162 vaccine reported a Grade 4 local reaction.

**Figure 2.**
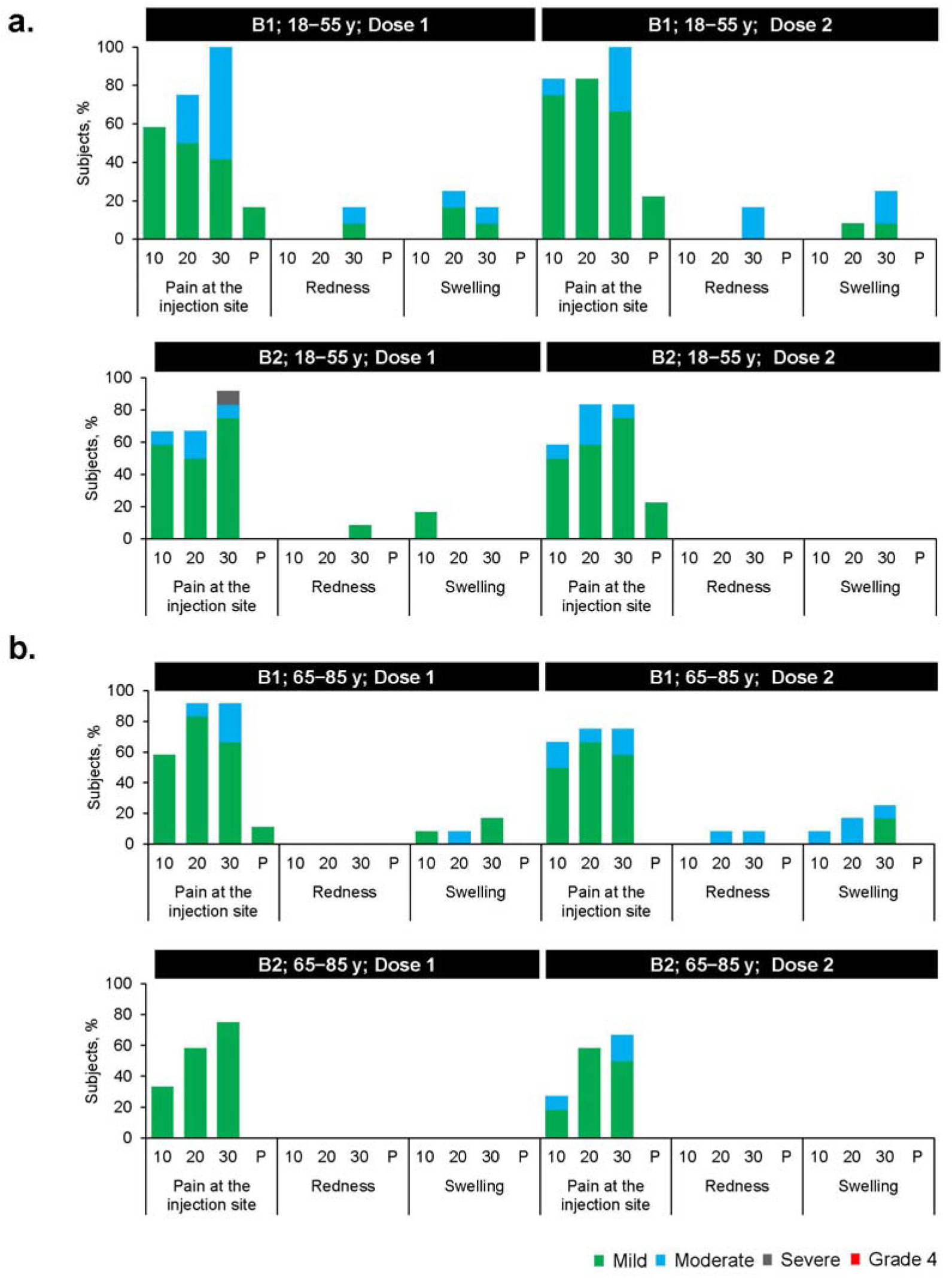
| Local reactions reported within 7 days after vaccination by age group (a. 18–55 years of age; b. 65–85 years of age). Solicited injection-site (local) reactions were collected with electronic diaries for 7 days after each vaccination. Pain at injection site scale – mild: does not interfere with activity; moderate: interferes with activity; severe: prevents daily activity; Grade 4: emergency room visit or hospitalization. Redness and swelling scale – mild: 2.0 to 5.0 cm in diameter; moderate: >5.0 to 10.0 cm in diameter; severe: >10.0 cm in diameter; Grade 4: necrosis or exfoliative dermatitis for redness, and necrosis for swelling. 10 = 10 μg; 20 = 20 μg; 30 = 30 μg; P = placebo; B1 – BNT162b1; B2 – BNT162b2.

##### Systemic events

Participants 18–55 years old who received 10 or 30 μg BNT162b1 frequently experienced mild to moderate fever and chills, with 75% reporting fever >38.0°C after Dose 2 of 30 μg (Figure 3 and Figure S1).^5^ In 65–85 year old participants who received BNT162b1, systemic events were milder than in the younger participants, though many older participants reported fatigue and headache after Dose 1 or Dose 2, and 33% of older participants reported fever >38°C after Dose 2, including 1 older participant who reported fever (38.9–40.0°C; Figure 3 and Figure S2). Like local reactions, systemic events were dose-dependent, greater after Dose 2 than Dose 1, and transient. Symptoms generally peaked by Day 2 after vaccination and resolved by Day 7.

**Figure 3.**
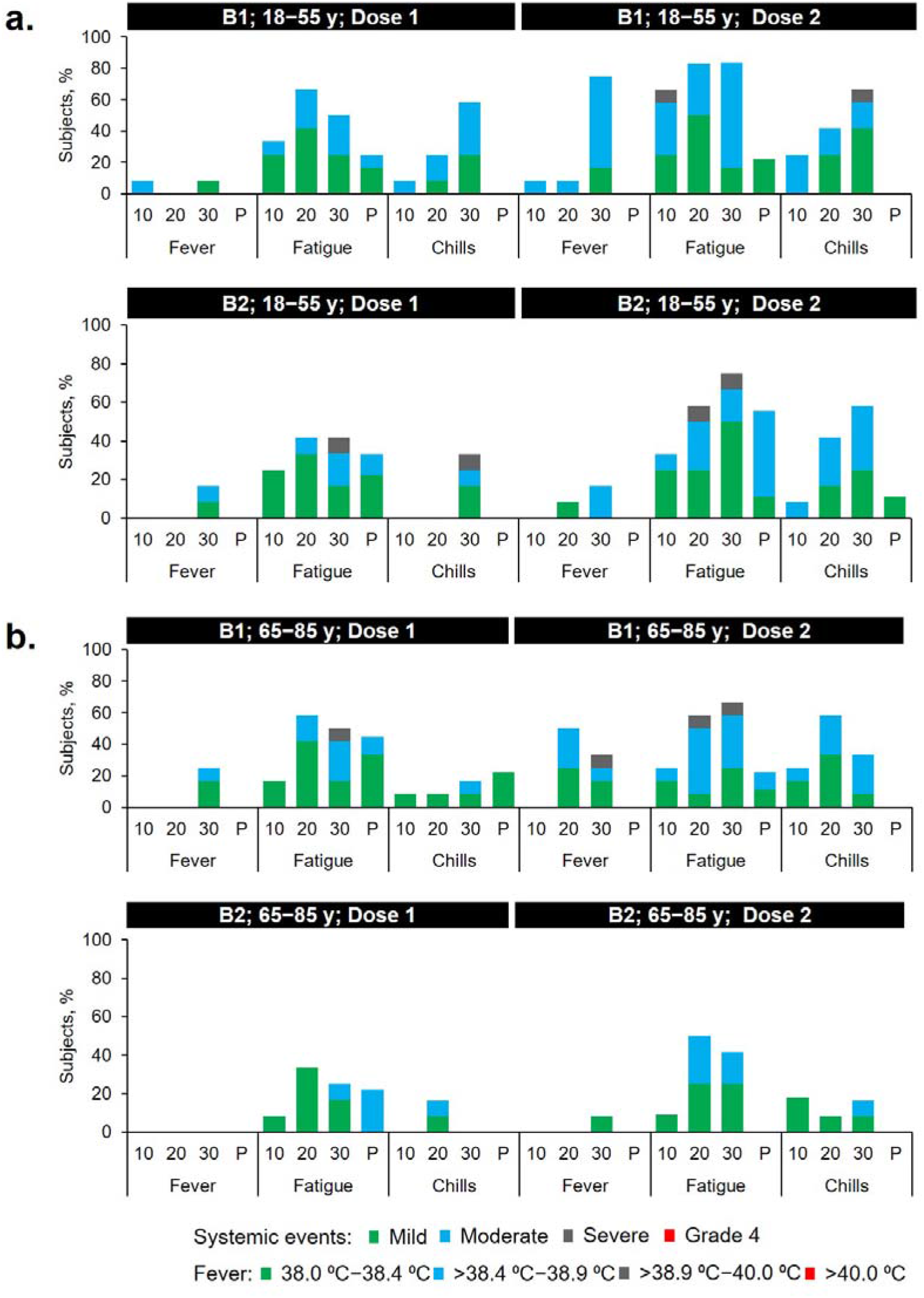
| Select systemic events reported within 7 days after vaccination (a. 18–55 years of age; b. 65–85 years of age). Fever, chills, and fatigue reported here. Headache, vomiting, diarrhea, muscle pain, and joint pain reported in Figure S1. Systemic events were collected with electronic diaries for 7 days after each vaccination. Fever scale as indicated in the key. Chills and fatigue scale – mild: does not interfere with activity; moderate: some interference with activity; severe: prevents daily activity; Grade 4: emergency room visit or hospitalization. 10 = 10 μg; 20 = 20 μg; 30 = 30 μg; P = placebo; B1 – BNT162b1; B2 – BNT162b2.

Systemic events in response to BNT162b2 were milder than those to BNT162b1 (Figures 3, S1, and S2). For example, only 17% of 18–55 year olds and 8% of 65–85 year olds reported fever (>38.0–38.9°C) after Dose 2 of 30 μg BNT162b2. Severe systemic events (fatigue, headache, chills, muscle pain, and joint pain) were reported in small numbers of younger BNT162b2 recipients, but no severe systemic events were reported by older BNT162b2 recipients. There were no reports of Grade 4 systemic events by any BNT162 recipient. Overall, systemic events reported by 65–85 year olds who received BNT162b2 were similar to those reported by those who received placebo after Dose 1.

#### Adverse Events and Shifts in Laboratory Values

Through 1 month after Dose 2, 50% of 18–55 year old participants who received 30 μg BNT162b1 reported related AEs compared to 11.1% of placebo recipients.^5^ Among 65–85 year olds who received 30 μg BNT162b1 and 18–55 year olds who received 30 μg BNT162b2, 16.7% reported related AEs (Table S1). No 65–85 year old who received 30 μg BNT162b2 reported a related AE. No SAEs were reported, and no stopping rules were met as of the time of this report. The largest changes from baseline in laboratory values were transient decreases in lymphocyte counts, which resolved within a week after vaccination (Figure S3) and were not associated with clinical manifestations.

### Immunogenicity

The serological responses elicited by BNT162b1 and BNT162b2 were similar. Antigen-binding IgG and neutralizing responses to vaccination with 10 μg to 30 μg of BNT162b1 or BNT162b2 were boosted by Dose 2 in both younger^2,5^ and older adults, showing clear benefit of a second dose (Figure 4a). Both vaccines elicited lower antigen-binding IgG and neutralizing responses in 65–85 year olds compared to 18–55 year olds (Figure 4). For example, neutralizing GMTs at Day 28 (7 days post Dose 2) of older adults who received 30 μg of BNT162b1 or BNT162b2 were 0.38 and 0.41 times, respectively, the GMTs of the corresponding younger adult cohorts (Figure 4b). Although there was a clear dose-level response between 10 μg and 20 μg of either vaccine candidate, a dose-level response between 20 μg and 30 μg was not consistent by vaccine candidate or by age group (Figure 4). Neutralizing GMTs measured 7 days after Dose 2 of 30 μg BNT162b1 or BNT162b2 ranged from 1.1 to 1.6 times the convalescent serum panel GMT in 65–85 year olds and from 2.8 to 3.8 times the convalescent serum panel GMT in 18–55 year olds.

**Figure 4.**
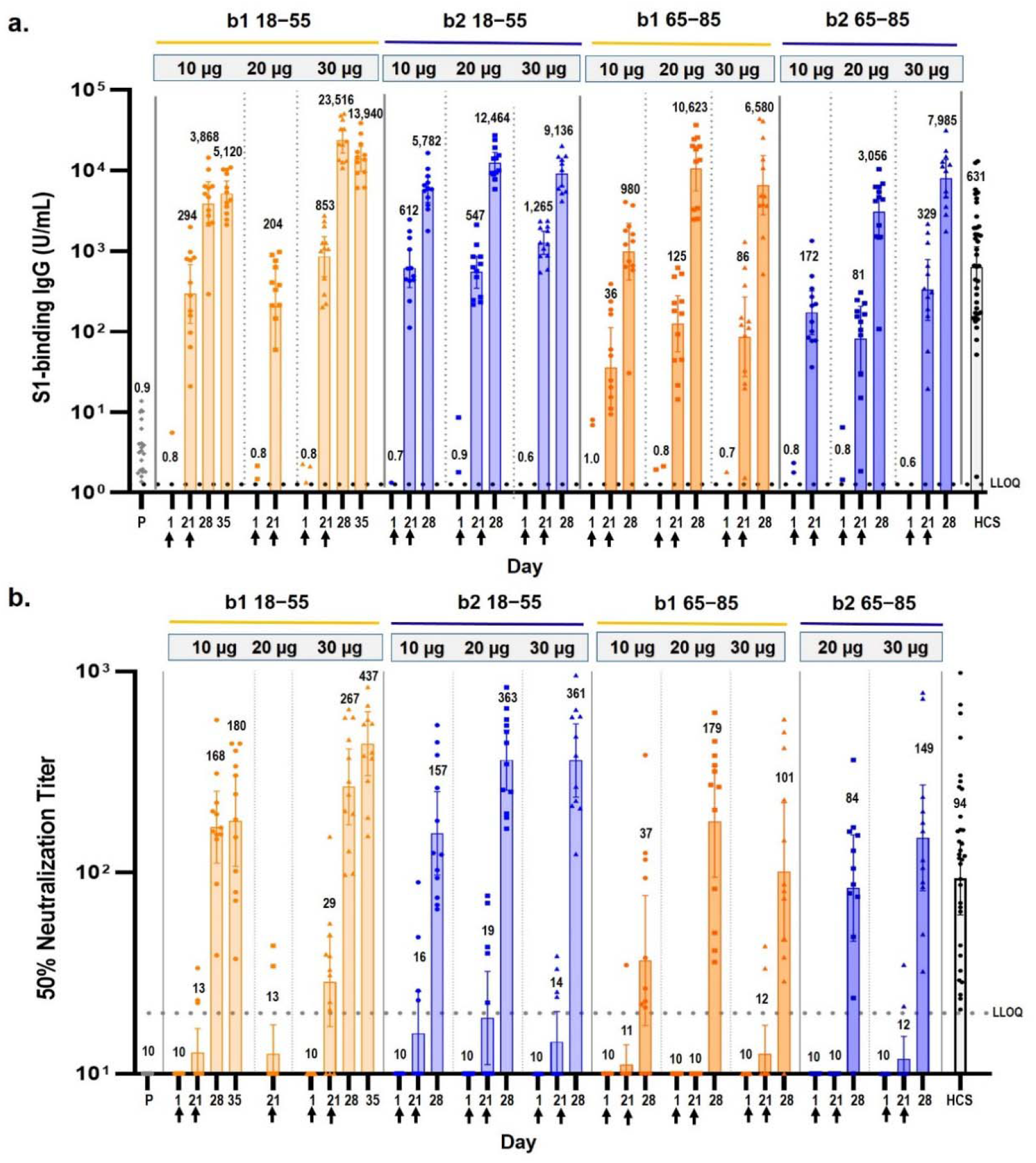
| Immunogenicity of BNT162b1 and BNT162b2. Participants in groups of 15 were vaccinated with the indicated dose levels of either BNT162b vaccine candidate (n=12) or with placebo (n=3) on Days 1 and 21. Reponses in placebo recipients for each of the dosing groups are combined (P). The 28-day blood collection is 7 days after the second vaccination. Sera were obtained before vaccination (Day 1) and 21, and 28 days after the first vaccination. Day 35 is shown only for the 10 μg and 30 μg BNT162b1 (18–55 years) groups. Human COVID-19 convalescent sera (HCS, n=38) were obtained at least 14 days after polymerase chain reaction- confirmed diagnosis and at a time when the donors were asymptomatic. a. Geometric mean concentrations (GMCs) of recombinant S1-binding IgG. Lower limit of quantitation (LLOQ) is 1.267. b. 50% SARS-CoV-2-neutralizing geometric mean titers (GMTs). LLOQ is 20. Each data point represents a serum sample, and each vertical bar represents a geometric mean with 95% CI. The numbers above the bars are the GMCs or GMTs for the group. Arrows indicate timing of vaccination (blood draws were conducted prior to vaccination on vaccination days).

## DISCUSSION

Previously reported data from vaccination of 18–55 year old adults with 10 μg or 30 μg of BNT162b1 suggested that it could be a promising COVID-19 vaccine candidate.^2,5^ Consistent with our strategy to evaluate several RNA vaccine candidates and make a data-driven decision to advance the candidate with the best safety and immunogenicity profile, we compared clinical data obtained after vaccination with BNT162b1,^2,5^ which encodes the RBD, or with BNT162b2, which expresses the full-length spike. The data set presented here guided our decision to advance BNT162b2 at the 30-μg dose level into the Phase 2/3, global safety and efficacy evaluation in participants 18–85 years of age.

The primary consideration driving this decision was the milder systemic reactogenicity profile of BNT162b2, particularly in older adults, in the context of comparable antibody responses elicited by both candidate vaccines. Short-lived declines in postvaccination lymphocyte counts were without evidence of associated clinical impact, were observed across age groups, and likely reflect temporary redistribution of lymphocytes from the bloodstream to lymphoid tissues as a functional response to the immune stimulation of immunization.^13,14,15,16^ The observation of a modRNA vaccine candidate at the selected, relatively low dose level of 30 μg that is both very immunogenic and well tolerated is unexpected for a modRNA vaccine candidate targeting an infectious disease.^17,18^ Lipid composition of the LNPs, formulation components or sequence selection for the RNA backbone and/or antigen target could influence the tolerability profile. The reason for the lower reactogenicity of BNT162b2 compared to BNT162b1 is not certain, given that BNT162b1 and BNT162b2 share the same modRNA platform, RNA production and purification processes, and LNP formulation. They differ in the nucleotide sequences encoding the vaccine antigens and in the overall size of the RNA constructs, resulting in approximately five times the number of RNA molecules in 30 μg of BNT162b1 compared to 30 μg of BNT162b2. The nucleotide composition of RNA appears to affect its immune stimulatory activity and reactogenicity profile.^19^

The immune responses elicited by BNT162b1 and BNT162b2 were similar. As observed with other vaccines and likely associated with immunosenescence,^20,21^ the immunogenicity of both vaccine candidates decreased with age, eliciting lower humoral responses in 65–85 year olds than in 18–55 year olds. Nevertheless, at 7 days after Dose 2, the neutralizing GMT elicited by 30 μg BNT162b2 in older adults, despite being only 0.41 times the GMT of younger adults, still exceeded the GMT of the convalescent serum panel. Based on the responses to 30 μg of BNT162b1 (Figure 4b), neutralizing GMTs elicited by BNT162b2 are expected to further increase from 7 to 14 days after Dose 2 (Day 35). At this time point, the GMT elicited by BNT162b1 was 4.6 times the GMT of the convalescent serum panel.^5^

A subtle difference in the humoral response to BNT162b1 and BNT162b2 is an apparent dose-response plateau for both vaccines in younger adults, but only for BNT162b1 in older adults. The more reactogenic regimens are associated with an apparent dose-response plateau. RNA vaccines require vaccine RNA translation in the host to express antigen, thus higher reactogenicity may be associated with an innate immune shutdown of host cell translation that can result in suboptimal antigen presentation and lower immunogenicity.

This study and interim report have several limitations. First, at the time of publication, data on immune responses or safety beyond 7 days after Dose 2 were not available. Second, we do not yet know the relative importance of humoral and cellular immunity in protection from COVID-19. Although strong cell-mediated immune responses (T_H_1-biased CD4^+^ and CD8^+^) elicited by BNT162b1 have been observed and reported from the German trial,^2^ cellular immune responses elicited by BNT162b2 are still being studied and will be reported separately. We anticipate that the full-length spike encoded by BNT162b2 will present a greater diversity of T-cell epitopes than does the much smaller RBD encoded by BNT162b1. This may lead to stronger and more consistent cellular responses to BNT162b2. Third, although the serum neutralizing responses elicited by the vaccine candidates relative to those elicited by natural infection are highly encouraging, the degree of protection against COVID-19 provided by this or any other benchmark is unknown. Finally, participants in this early-stage clinical study were healthy and in groups too small to reflect the diversity of those in need of a COVID-19 vaccine.

Many of the limitations to this study are now being addressed in the global Phase 2/3 portion of this study, while we expand our RNA vaccine manufacturing and distribution capacity. In this pivotal study, we are assessing the safety and efficacy of 2 doses of 30 μg BNT162b2 in up to 30,000 participants (randomized 1:1 with placebo) from diverse backgrounds, including individuals with stable chronic underlying health conditions, individuals at increased risk due to occupational exposure, and individuals from racial and ethnic backgrounds at higher risk for severe COVID-19.^22^

## Data Availability

Upon request, and subject to review, Pfizer will provide the data that support the findings of this study. Subject to certain criteria, conditions, and exceptions, Pfizer may also provide access to the related individual anonymized participant data.

https://www.pfizer.com/science/clinical-trials/trial-data-and-results

## Role of the funding source

BioNTech is the sponsor of the study. Pfizer was responsible for the design, data collection, data analysis, data interpretation, and writing of the report. The corresponding authors had full access to all the data in the study and had final responsibility for the decision to submit the data for publication. All study data were available to all authors.

## Disclosure

These data are interim data from an ongoing study, with the database not locked. Data have not yet been source verified or subjected to standard quality check procedures that would occur at the time of database lock and may therefore be subject to change.

## Data sharing statement

Upon request, and subject to review, Pfizer will provide the data that support the findings of this study. Subject to certain criteria, conditions, and exceptions, Pfizer may also provide access to the related individual anonymized participant data. See https://www.pfizer.com/science/clinical-trials/trial-data-and-results for more information.

## Acknowledgements

The authors would like to thank Carol Monahan and Deb Gantt (Pfizer Inc) for writing and editorial support, James Trammel (Pfizer Inc) for statistical analysis support in the generation of this manuscript, and Tricia Newell (ICON plc, North Wales, PA) for editorial support.

We would like to thank all the participants who volunteered for this study. We also acknowledge the following individuals for their contributions to this work:

NYU Langone Vaccine Center: Angelica Kottkamp, MD, Ramin Herati, MD, Rebecca Pellet Madan, MD, Mary Olson, DNP, ANP-BC, Marie Samanovic-Golden, PhD, Elisabeth Cohen, MD, Amber Cornelius, MS, Laura Frye, MPH, Heekoung Youn, RN, CCRC, MA, Baby Jane Fran, RN, Kanika Ballani, PharmD, MBA, Natalie Veling, RN, Juanita Erb, RN, BSN, MPA, Mahnoor Ali, BA, Lisa Zhao, BA, Stephanie Rettig, MPH, Hibah Khan, MPA, Harry Lambert, BA, Kelly Hu, BA, and Jonathan Hyde, BS. Staffing services were supported in part by an NYU CTSA grant (UL1 TR001445) from the National Center for Advancing Translational Sciences, National Institutes of Health.

Center for Vaccine Development and Global Health, University of Maryland School of Medicine: Monica McArthur, MD, PhD, Justin Ortiz, MD, MS, FACP, FCCP, Rekha Rapaka, MD, Linda Wadsworth, RN, Ginny Cummings, RN, Toni Robinson, RN, Nancy Greenberg, RN, Lisa Chrisley, RN, Wanda Somrajit, RN, Jennifer Marron, RN, BSN, MS, Constance Thomas, RN, Kelly Brooks, RN, Lisa Turek, RN, Patricia Farley, RN, Staci Eddington, Panagiota Komninou, Mardi Reymann, Kathy Strauss, Biraj Shrestha, Sudhaunshu Joshi, Robin Barnes, RN, Roohali Sukhavasi, Myounghee Lee, PharmD, Alyson Kwon, and Terry Sharp.

University of Rochester and Rochester General Hospital: Emily Pierce, RN, Mary Criddle, RN, Maryrose Laguio-Vila, MD, Megan Helf, MS, Madison Murphy BS, Maria Formica, MS, and Sarah Korones, MD.

Cincinnati Children’s Hospital: Amy Cline, RN, Susan Parker, RN, and Michelle Dickey,

APRN, Kristen Buschle, APRN.

Pfizer Inc: Andrea Cawein, John L. Perez, MD, MSc, Harpreet Seehra, Dina Tresnan, DVM, PhD, Robert Maroko, MD, Helen Smith, Sarah Tweedy, Amy Jones, Greg Adams, Rabia Malick, Emily Worobetz, Erica Weaver, Liping Zhang, Carmel Devlin, Donna Boyce, Elisa Harkins Tull, Mark Boaz, Michael Cruz, Vaccines Clinical Assay Team, and Vaccines Assay Development Team.

BioNTech: Corinna Rosenbaum, Christian Miculka, Andreas Kuhn, Ferdia Bates, Paul Strecker, and Alexandra Kemmer-Brück.

## SUPPLEMENTARY APPENDIX

**Figure S1.**
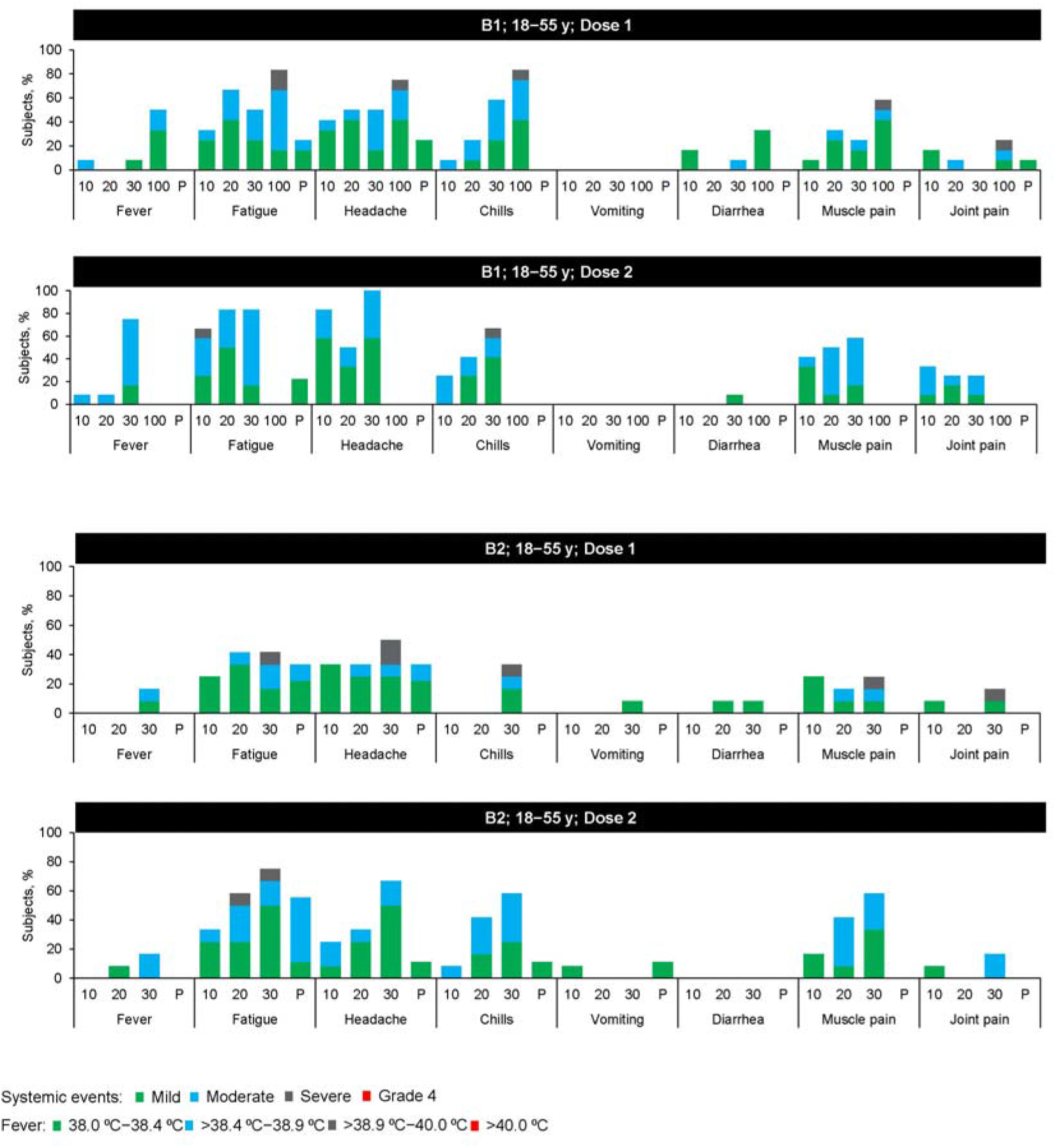
| Systemic events reported within 7 days after vaccination, 18–55 years of age. Systemic events were collected with electronic diaries for 7 days after each vaccination. Fever scale as indicated in the key. Fatigue, headache, chills, new or worsened muscle pain, new or worsened joint pain (mild: does not interfere with activity; moderate: some interference with activity; severe: prevents daily activity), vomiting (mild: 1 to 2 times in 24 hours; moderate: >2 times in 24 hours; severe: requires intravenous hydration) and diarrhea (mild: 2 to 3 loose stools in 24 hours; moderate: 4 to 5 loose stools in 24 hours; severe: 6 or more loose stools in 24 hours); Grade 4 for all events: emergency room visit or hospitalization; 10 = 10 μg; 20 = 20 μg; 30 = 30 μg; P = placebo; B1 – BNT162b1; B2 – BNT162b2. A second dose of BNT162b1 100 μg was not given to participants in the 18–55 year old group because of unsatisfactory tolerability after the first dose.

**Figure 2.**
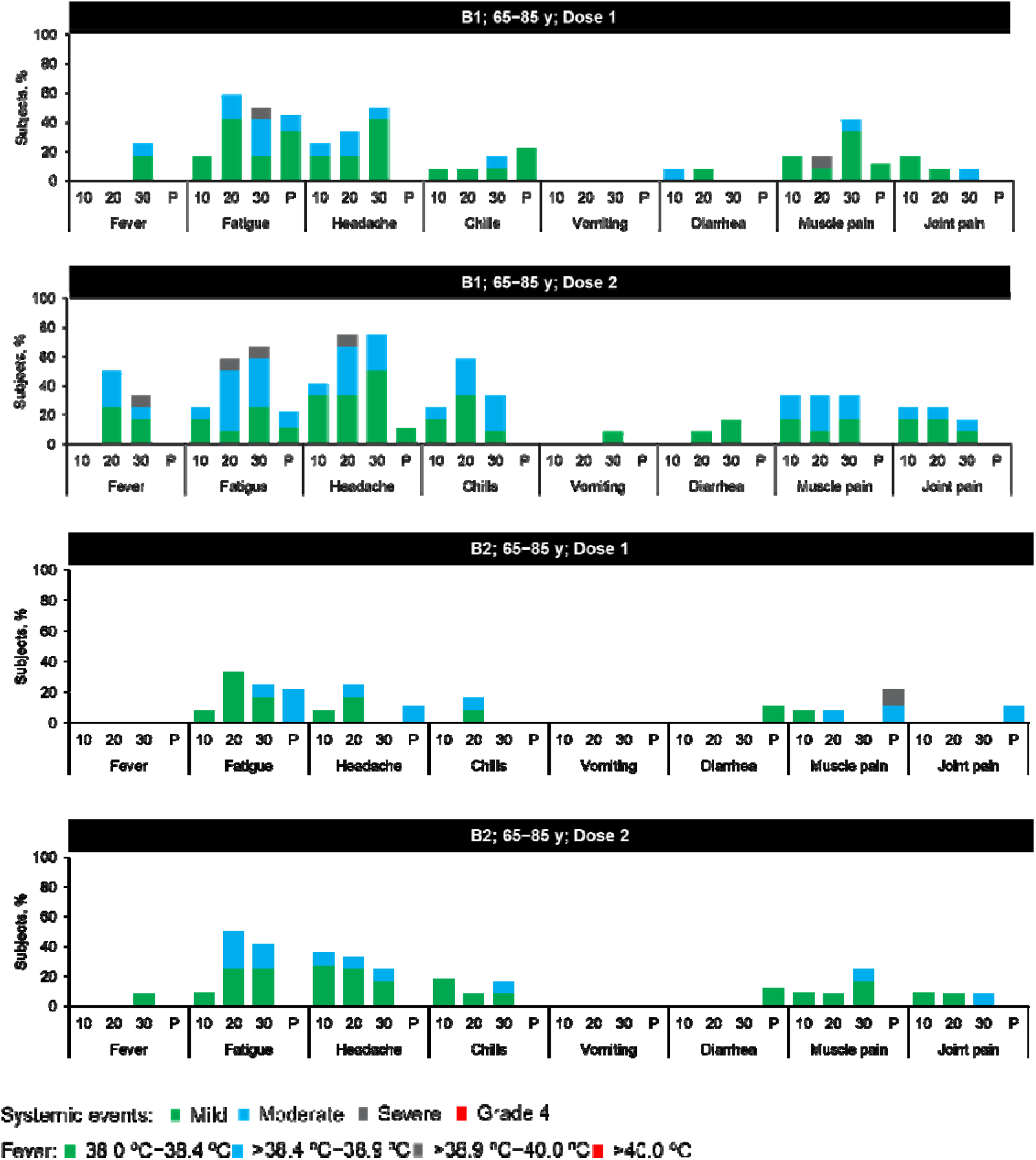
| Systemic events reported within 7 days after vaccination, 65–85 years of age. Systemic events were collected with electronic diaries for 7 days after each vaccination. Fever scale as indicated in the key. Fatigue, headache, chills, new or worsened muscle pain, new or worsened joint pain (mild: does not interfere with activity; moderate: some interference with activity; severe: prevents daily activity), vomiting (mild: 1 to 2 times in 24 hours; moderate: >2 times in 24 hours; severe: requires intravenous hydration) and diarrhea (mild: 2 to 3 loose stools in 24 hours; moderate: 4 to 5 loose stools in 24 hours; severe: 6 or more loose stools in 24 hours); Grade 4 for all events: emergency room visit or hospitalization; 10 = 10 μg; 20 = 20 μg; 30 = 30 μg; P = placebo; B1 – BNT162b1; B2 – BNT162b2. A second dose of BNT162b1 100 μg was not given to participants in the 18–55 year old group because of unsatisfactory tolerability after the first dose.

**Figure 3.**
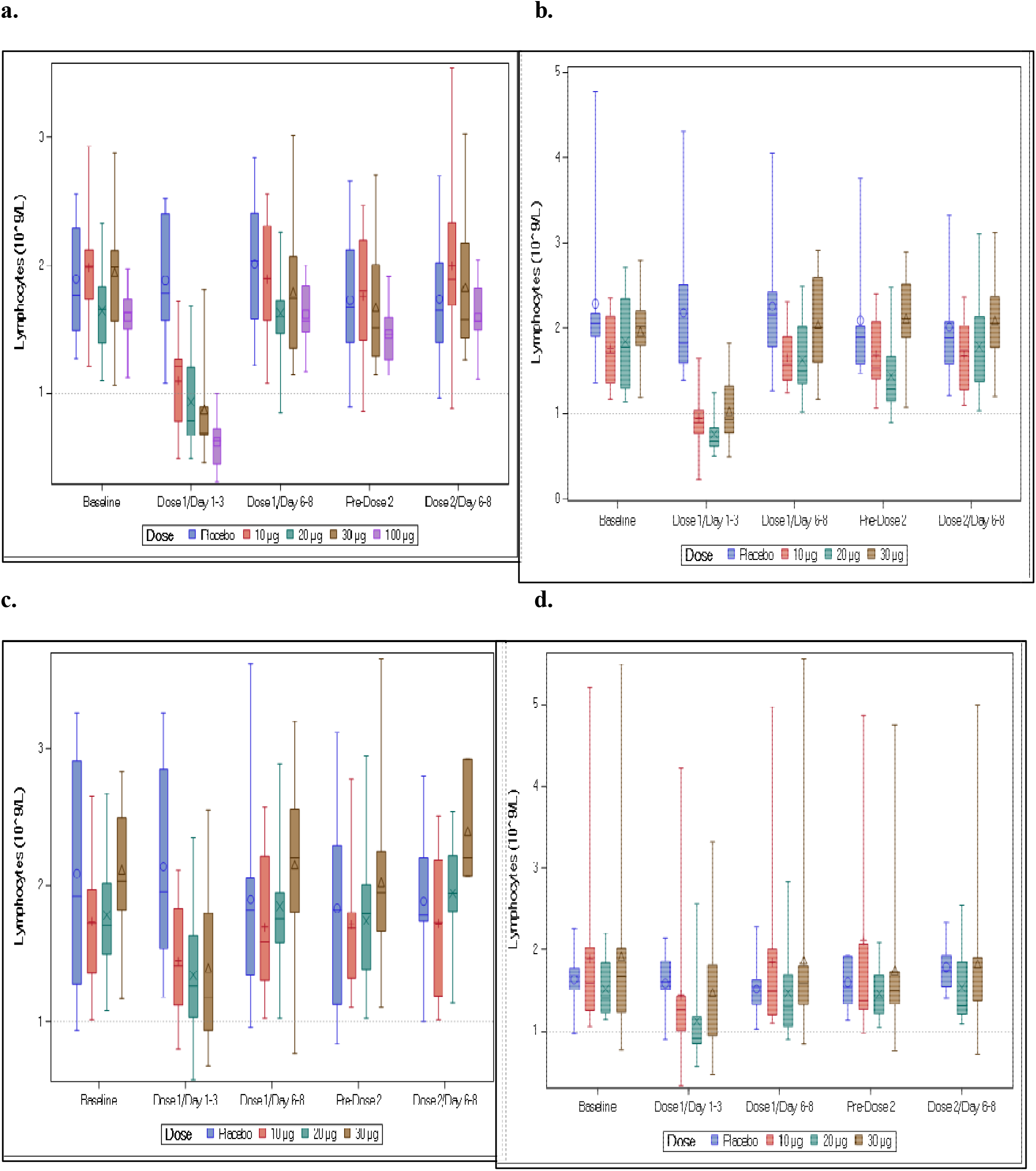
| Postvaccination changes in lymphocyte count over time. Figure represents box- and-whisker plots for observed values at the following time points: Dose 1/Day 1–3: ~1 day after Dose 1; Dose 2/Day 6–8: ~7 days μafter Dose 1; Pre-Dose 2: before Dose 2; Dose 2/Day 6–8: ~ 7 days after Dose 2. Symbols denote group means – O: placebo; **+:** 10 μg; X: 20 μg; Δ: 30 μg;: 100 μg. Center line of box denotes median; lower and upper edges denote first and third quartiles; lower and upper whiskers denote minimum and maximum. **a**. BNT162b12 18–55 years of age; **b**. BNTl62bl 65—85 years of age; **c**. BNTl62b2 18—55 years of age; **d**. BNTl62b2 65— 85 years of age.

**Table S1.**
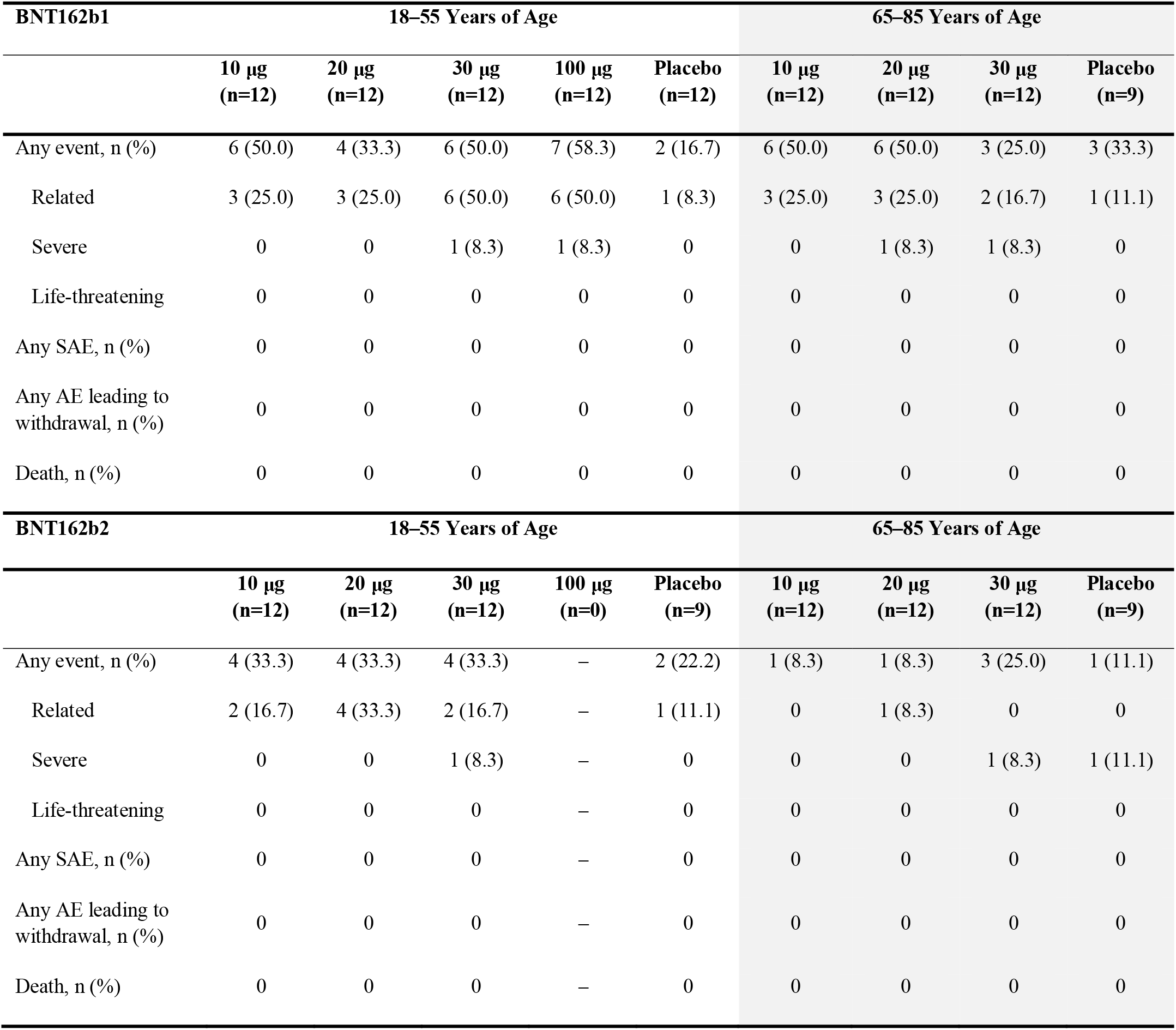
| Adverse events by age group and vaccine candidate. AE=adverse event. Related AE=adverse event that in the opinion of the investigator was possibly related to study vaccine. SAE=serious adverse event.

## Notes

### Competing Interest Statement

Competing interests: NK, JA, AG, SL, RB, KAS, PL, KK, WK, DC, KRT, PRD, WCG, and KUJ are employees of Pfizer and may hold stock options. US and OT are stock owners, management board members, and employees at BioNTech SE (Mainz, Germany) and are inventors on patents and patent applications related to RNA technology.  MJM, KEL, KN, EEW, ARF, RF, and VR received compensation from Pfizer for their role as study investigators.  CFG and PYS received compensation from Pfizer to perform the neutralization assay.

### Clinical Trial

NCT04368728

### Author Declarations

The study was approved by the institutional review boards for each participating site prior to enrollment of any study participant in this study

### Summary of Updates

Minor corrections to Table 1, Fig 1a and Fig S1

